# Spatial and Temporal Patterns of Community Utilization of a Tribally Run Shellfish Toxin Testing Program in the Gulf of Alaska

**DOI:** 10.1101/2025.07.01.25330576

**Authors:** Esther G. Kennedy, John R. Harley, Shannon M. Cellan, Kari Lanphier, Jeff Feldpausch, Christopher Whitehead, Matthew O. Gribble, Hugh B. Roland

## Abstract

Rural shellfish harvesters, including many Alaska Native peoples, require safe access to wild shellfish for subsistence, food security, and cultural practices. However, wild shellfish may be contaminated with paralytic shellfish toxins, leaving harvesters with increased risks of significant illness or death. To manage these risks, the Sitka Tribe of Alaska Environmental Research Lab (STAERL) was established to test shellfish samples sent in by harvesters in the community and to support regular monitoring of select local beaches by tribal governments. Here, we investigated harvester utilization of this shellfish testing service from 2016-2024, comprising 299 samples sent in by local harvesters, and used generalized linear models to examine how annual testing rates varied by year, location, species, and species-based detoxification rates. We pay particular attention to differences that may reflect the influence of risk perceptions and accessibility of harvesting and testing on utilization (DOI: 10.5061/dryad.dfn2z35dr). We find that testing utilization has increased through time (1.278, 95% CI: 1.161, 1.407), testing rates are highest in spring and broadly similar between the other three seasons, testing rates in Sitka are much higher than those outside of it, and neither road accessibility nor species-based detoxification rates strongly affect testing rate ratios. These findings suggest that shellfish testing behavior is consistent despite seasonal variations in risk and convenience, but that the STAERL individual testing program provides a pathway to maintain established subsistence harvest practices while reducing poisoning risks.

## 1. BACKGROUND

Coastal peoples worldwide harvest shellfish for traditional uses and subsistence (1–3). In rural communities, high food costs and limited access to stores make subsistence harvest an important practice for food security, in addition to its cultural importance (4,5). Though shellfish can serve as a reliable, healthy source of protein and other nutrients, they can also concentrate toxins produced by harmful algal blooms, often referred to as “red tides”, which can lead to deadly illnesses such as paralytic shellfish poisoning (PSP) (6,7). In some regions, risk of PSP may rise with climate change, as warming waters may contribute to greater frequency and intensity of harmful algal blooms and subsequently higher toxin concentrations in shellfish (8–10). The presence of shellfish toxins, which cannot be tasted or rendered inert by cooking or freezing, can suppress harvesting practices (11) or, in some cases, lead to widespread serious illness or death (12–14).

The challenge of shellfish toxicity has engendered many traditional harvesting practices such as avoiding certain tissues or times of year (11,15–17). In much of the coastal U.S., the threat of shellfish toxicity has also produced state-run toxin testing programs to mitigate those risks (18,19). In rural Alaska, however, shellfish harvesters must balance strong cultural and socioeconomic incentives to harvest without the support of a state-run toxin monitoring system for non-commercial users (15,20), while facing documented increases in algal bloom frequency (Anderson et al., 2021, p. 202; Yan et al., 2020) as well as a recent history of impactful and deadly PSP outbreaks in the region (12,14,21). In Southeast Alaska, tribal governments have developed a testing and monitoring program to facilitate and support safe shellfish harvests that is open to harvesters throughout the state, ensuring continued access to traditional resources in a changing environment (22).

### 1.1 Traditional shellfish harvesting practices in Southeast Alaska

Present-day Southeast Alaska is located within the traditional territories of the Tlingit, Haida, and Tsimshian people. The region faces pressing environmental, sociocultural, and health issues. Changing climate conditions threaten traditional resources such as yellow cedar (23) sockeye salmon (24,25) and shellfish (26). Shellfish harvesting has been widely practiced in the region for millennia (27,28), appears as a motif in traditional artwork (29), and is part of traditional food education (30,31). In the most recent surveys of household shellfish use by the Alaska Department of Fish and Game, rates of shellfish consumption varied substantially by community, ranging from 17% in Sitka to above 30% in Petersburg, Wrangell, and Yakutat and above 50% in Hoonah (32). Butter clams are the most heavily harvested species, with average per capita harvests of 4.36 pounds in 2012 in Southeast Alaska and 9.2 pounds in Kodiak in 2018, while cockles, littleneck clams, and scallops are also commonly harvested (32). While longitudinal data for most communities are sparse, harvest rates in many communities have been declining through time (32), in part because harvesters fear shellfish toxins (11,15).

### 1.2 Tribally led management of toxin risks

Culturally central consumption puts Tlingit, Haida, Tsimshian, and other coastal Alaska Native people at elevated risk of toxin exposure, particularly as warming waters undermine traditional seasonal strategies for reducing risks (11,15,22). Consumption of shellfish with high toxin concentrations may lead to serious health consequences, including dizziness, nausea, vomiting, and death, and coastal Alaska Natives’ traditional harvesting practices have contributed to wide health disparities (13). Fifty-three percent of paralytic shellfish poisoning (PSP) cases reported in Alaska between 1993-2021 were in Alaska Natives, despite Alaska Natives only making up 16% of Alaska’s population (14). In two coastal Alaska communities, Alaska Natives were 11.6 times as likely to report a history of PSP than non-Natives, with 20% of Alaska Natives reporting a history of symptoms (33). Butter clams are the most commonly implicated species in PSP events as they can retain high toxin levels for months or years and are frequently harvested (15,34,35), but mussels and cockles, also commonly harvested for subsistence, also contribute to Alaskan PSP cases (12,13).

To address disproportionate exposure risks and health disparities as well as to facilitate safe traditional harvesting practices, tribal governments across Southeast Alaska initiated a tribally led shellfish toxin testing program, forming the Southeast Alaska Tribal Ocean Research consortium (SEATOR) to serve as a regional partnership for environmental research and toxin testing and provide near real-time toxin data. The shellfish testing program involves routine sampling and testing of shellfish for toxins at specific, commonly used subsistence harvesting sites in participating Southeast Alaska communities. Each SEATOR partner government regularly collects shellfish samples at one or more community harvest sites and ships samples to the Sitka Tribe of Alaska Environmental Research Lab (STEARL) in Sitka, Alaska for toxin testing. Species analyzed by each community reflect the availability, but in general four species are targeted for routine testing regardless of season – mussels (Mytilus complex), cockles (Clinocardium nuttallii), butter clams (Saxidomus gigantea), and littleneck clams (Leukoma staminea). Toxin concentrations are reported back to partner communities on the web and via email, and concentrations exceeding safety thresholds are further disseminated as advisories through radio, social media, and physical notices. Recent research demonstrates evidence of the toxin testing program’s efficacy and community environmental managers report that harvesters increasingly check the SEATOR website for toxin levels before harvesting (11,22).

In addition to testing samples regularly collected by environmental managers in the SEATOR consortium, STAERL tests ad hoc samples sent by individual harvesters, originally charging $50/sample but making the service free in 2021. This service enables community members to test shellfish harvested from unmonitored locations prior to consumption. The toxin levels from ad hoc shellfish tests are only shared with the harvester rather than made public, as many harvesters prefer to keep their harvesting beds secret; however, STAERL issues a broad area-wide advisory if these opportunistic samples are above FDA-approved toxin levels, balancing harvester confidentiality and community safety. State-run shellfish toxin testing programs in other states have been effective in reducing exposure risks (18,19), but few shellfish toxin testing programs are community led, aim to inform subsistence harvesting decisions, and provide options for individual participation. Community-based risk management like SEATOR’s testing improve communities’ access to information and resources and increase local engagement and agency in environmental and health protection (36,37). The additional option for individual shellfish assessments through STAERL provides harvesters with the opportunity to assess risk for rare or regionally specific species, remote or culturally important locations, and specific timing windows that may be missed by bimonthly monitoring.

We aim to better understand utilization of the personal shellfish testing program as harvesters seek safe access to shellfish beds that are not regularly monitored, particularly the possible influences of risk perception and site and testing accessibility. Reviews examining factors contributing to public participation in climate adaptation strategies note widespread calls for increased community participation and a focus on behavioral factors and risk perceptions as factors shaping participation (38). For example, studies have examined risk perceptions as a barrier to participation in state-led lead exposure risk reduction strategies (39) and risk perceptions and perceived ability to reduce risks as barriers to participation in community-based disaster risk management (40). More limited research has examined barriers related to access to adaptation strategies, particularly in contexts of community-run adaptation (11). Here, we test three utilization hypotheses: 1) community utilization of the shellfish testing lab increases through time, possibly related to increased familiarity with the program, 2) testing rates will be highest in the spring and summer, when algal blooms are most common, and, further, that species with fast (slow) detoxification rates will be especially tested in the spring (winter) when risk conditions are changing the fastest (slowest), possibly related to harvesters’ perceptions of risk, and 3) more difficult to access harvesting sites, such as those inaccessible by road, and greater effort required to get samples to STAERL will reduce program utilization. By considering factors possibly related to risk perception and adaptation accessibility, this research may enhance understanding of factors contributing to community participation in risk reduction strategies and inform efforts to increase utilization of diverse environmental management approaches.

## 2. DATA AND METHODS

### 2.1 Data sources

We evaluate individual, ad-hoc shellfish samples sent to STEARL for testing by community members from the start of the program in 2016 through 2024. The shellfish testing data provided to researchers included the date samples were collected, the date samples were analyzed, reported harvesting site as described by the harvester, community from which the sample was sent, shellfish species, and tested paralytic shellfish toxin level (41). To preserve harvester confidentiality, the data shared did not include harvester identifiers so we could not assess whether repeated samples from a community reflected multiple individual harvesters or repeated harvests from a few individuals.

To adjust the number of samples sent by the population of the sending area, we used annual population data from the U.S. Census Bureau for each Census Area or borough that sent individual samples to STAERL during our study period (Figure 1; Table 1; (42)). For each Census Area or borough, we used the reported population size in 2020 and the census-provided projections for 2021-2024 and extrapolated 2016-2019 populations using the 2020-2024 growth rate. Finally, we grouped the populations into Sitka and non-Sitka groups, where the non-Sitka group comprised the combined populations of all of the non-Sitka boroughs that sent any individual shellfish over our study period.

**Figure 1:**
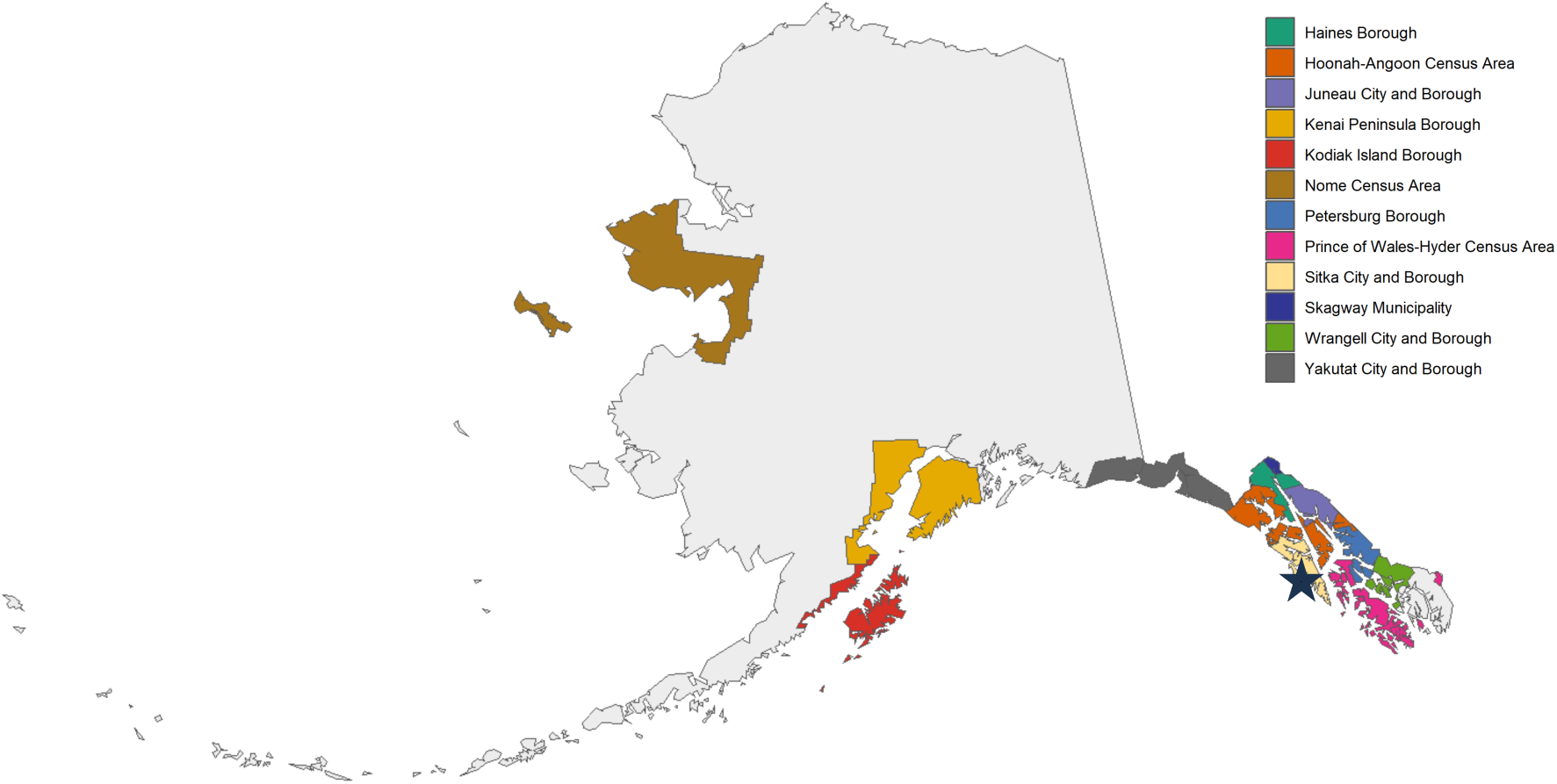
Map of the Alaskan boroughs and census areas that submitted individual shellfish samples to the Sitka Tribe of Alaska Environmental Research Lab (STAERL) for toxin testing from 2016 to 2024. The location of STAERL is marked by a black star.

**Table 1:** Borough population by year. Populations by borough or Census Area from the 2020 census. The 2021-2024 population levels are the census-reported projections. We derived the 2016-2019 annual populations using the 2020-2024 census-reported growth rate.

| Borough | 2016 | 2017 | 2018 | 2019 | 2020 | 2021 | 2022 | 2023 | 2024 |
| --- | --- | --- | --- | --- | --- | --- | --- | --- | --- |
| Haines Borough | 1680 | 1772 | 1869 | 1972 | 2080 | 2621 | 2582 | 2539 | 2537 |
| Hoonah-Angoon Census Area | 2486 | 2455 | 2425 | 2395 | 2365 | 2358 | 2355 | 2305 | 2248 |
| Juneau City and Borough | 33087 | 32877 | 32668 | 32461 | 32255 | 32251 | 31860 | 31616 | 31436 |
| Kenai Peninsula Borough | 56316 | 56927 | 57544 | 58168 | 58799 | 59047 | 60006 | 61003 | 61350 |
| Kodiak Island Borough | 13645 | 13507 | 13370 | 13235 | 13101 | 12943 | 12848 | 12766 | 12570 |
| Nome Census Area | 10452 | 10349 | 10247 | 10146 | 10046 | 9714 | 9685 | 9644 | 9651 |
| Petersburg Borough | 3418 | 3413 | 3408 | 3403 | 3398 | 3375 | 3356 | 3371 | 3379 |
| Prince of Wales-Hyder Census Area | 5765 | 5762 | 5759 | 5756 | 5753 | 5752 | 5727 | 5799 | 5740 |
| Skagway Municipality | 1364 | 1332 | 1301 | 1270 | 1240 | 1205 | 1145 | 1127 | 1123 |
| Wrangell City and Borough | 2227 | 2202 | 2177 | 2152 | 2127 | 2103 | 2086 | 2044 | 2030 |
| Yakutat City and Borough | 686 | 680 | 674 | 668 | 662 | 699 | 674 | 680 | 637 |
| **Sitka** | **8864** | **8761** | **8659** | **8558** | **8458** | **8414** | **8347** | **8203** | **8063** |
| **Total non-Sitka** | **131126** | **131276** | **131442** | **131626** | **131826** | **132068** | **132324** | **132894** | **132701** |

### 2.2 Variables coding

From the raw shellfish harvest data, we created several additional variables (41). First, we assigned each sample a season according to the meteorological calendar (Dec - Feb = Winter; Mar - May = Spring; June - Aug = Summer; Sep - Nov = Fall; National Oceanic and Atmospheric Administration, 2023) because, compared to astronomical season, meteorological season better aligns with traditional harvest windows (e.g., most salmon are harvested Jun - Aug, most deer are harvested Sep – Nov) and the meteorological calendar’s monthly breaks better align with a commonly quoted traditional shellfish harvesting adage in Southeast Alaska that advises to only harvest in months that contain the letter “r” as these months (Sep - Apr) are cooler.

To evaluate shellfish and testing accessibility, we created two spatial variables: a “Sitka vs. non-Sitka” variable to account for the requirement to ship sample to the testing lab for non-Sitkans and a “road status” variable denoting whether the harvesting site was accessible by road from the sending community, inaccessible by road, or unreported, potentially reflecting how difficult it may be to collect samples. We assigned each sample a population size from our extrapolated census data, using the sample year and corresponding Sitka or non-Sitka status. Ideally, we would also have derived an “importance factor” reflecting the reliance of the sending community on each species of shellfish, as reported by the Community Subsistence Information Survey, as the importance of a given resource might interact with the importance of convenience for testing utilization. However, the recent (post-2000) subsistence survey data available does not include several communities that routinely sent shellfish samples, such as Douglas, Metlakatla, or Klawock, and cannot be meaningfully grouped across communities or extrapolated to unreported areas (32). As such, we consider differences in shellfish reliance when interpreting our model results, but do not directly assess consumption or use metrics.

To evaluate the influence of species on seasonal or overall risk perception, we created a new “detoxification rate” variable, classifying shellfish species as having fast, slow, or unknown detoxification rates according to Bricelj and Shumway (1998) (6,35). However, since the “slow” detoxification category was very heavily weighted toward butter clams and scallops (69% and 30%, respectively), we also opted to examine the three most commonly harvested species (butter clams, scallops, and cockles), grouping all other species “other” in a “lumped species” variable.

### 2.3 Statistical Analysis

For all analyses, we grouped samples by year and by location (Sitka vs. non-Sitka), then further grouped them by species, road status, detoxification rate, or other predictor variables to generate counts through time (Table 2). We used stacked bar charts to visualize trends in the number of community samples tested by sending community, species harvested, and accessibility of the harvesting site and to generate our hypotheses (e.g., Figure 1). Four out of 299 total individual samples were reported without any location information, all sent in 2017; these were excluded from analyses.

**Table 2:** Annual shellfish sample counts by category. Annual individual shellfish samples sent to STAERL for testing by species, season, region, and road status. These categories are non-exclusive. Counts derived from the raw data hosted at https://doi.org/10.5061/dryad.dfn2z35dr (Kennedy et al., 2025).

|  |  | 2016 | 2017 | 2018 | 2019 | 2020 | 2021 | 2022 | 2023 | 2024 | Total |
| --- | --- | --- | --- | --- | --- | --- | --- | --- | --- | --- | --- |
| Species | Butter Clam | 1 | 10 | 9 | 8 | 21 | 24 | 10 | 21 | 28 | 132 |
|  | Geoduck | 1 | 0 | 0 | 0 | 0 | 0 | 0 | 0 | 1 | 2 |
|  | Scallop | 3 | 2 | 2 | 0 | 9 | 11 | 12 | 2 | 16 | 57 |
|  | Cockle | 0 | 2 | 0 | 4 | 9 | 6 | 4 | 6 | 6 | 37 |
|  | Littleneck Clam | 0 | 1 | 0 | 2 | 2 | 4 | 2 | 3 | 6 | 20 |
|  | Mussel | 0 | 3 | 0 | 2 | 2 | 2 | 3 | 2 | 4 | 18 |
|  | Oyster | 0 | 1 | 1 | 1 | 0 | 0 | 0 | 0 | 1 | 4 |
|  | Other | 0 | 3 | 0 | 1 | 3 | 6 | 1 | 1 | 14 | 29 |
| Season | Fall | 4 | 1 | 0 | 1 | 10 | 6 | 4 | 10 | 9 | 45 |
|  | Winter | 1 | 6 | 2 | 1 | 5 | 8 | 14 | 7 | 9 | 53 |
|  | Spring | 0 | 10 | 5 | 8 | 20 | 24 | 6 | 13 | 47 | 133 |
|  | Summer | 0 | 5 | 5 | 8 | 11 | 15 | 8 | 5 | 11 | 68 |
| Region | Non-Sitka Areas | 1 | 12 | 2 | 15 | 12 | 25 | 13 | 12 | 23 | 115 |
|  | Sitka Region | 4 | 10 | 10 | 3 | 34 | 28 | 19 | 23 | 53 | 184 |
| Road Status | Unreported | 5 | 8 | 1 | 6 | 4 | 7 | 4 | 1 | 21 | 57 |
|  | Off | 0 | 12 | 10 | 3 | 33 | 22 | 13 | 11 | 27 | 131 |
|  | On | 0 | 2 | 1 | 9 | 9 | 24 | 15 | 23 | 28 | 111 |
| Annual Shellfish Total |  | 5 | 22 | 12 | 18 | 46 | 53 | 32 | 35 | 76 |  |

We then fit generalized linear models (GLMs) with a negative binomial distribution and log link function to identify the most important factors contributing to individual utilization of shellfish testing (Table 3, end of document). For each group of tested predictor variables, we fitted a “set” of three GLMs: one including all data with Sitka vs. non-Sitka included as a fixed effect, one limited to Sitka data only, and one limited to non-Sitka data only. Model Set 1 only included year as a predictor and the log-transformed annual population as an offset term. Model Set 2 added seasons to the previous two predictors. On all subsequent models, year, seasons, and the population offset were all included. Model Set 3 examined known road status (on or off) of the harvesting site, dropping 57 samples with indeterminable road accessibility. Model Sets 4-6 examined species and detoxification rate influences, featuring a binary predictor for butter clams, a four-level species category (butter clams, scallops, cockles, and “other”), and species-based detoxification rates (fast, slow, and unknown), respectively. Finally, Model Set 7 included road status (on, off, or unreported), a binary species predictor (butter clams or not), and season. For Model Sets 3-7, we also assessed the evidence for interaction terms between season and species-based, road status, and/or year terms by using the likelihood-ratio test to compare models with interaction terms to their nested counterparts, with a ∝ of 0.05. We found no evidence for including interaction terms in any of these models and, as such, present only the nested models here.

**Table 3:**
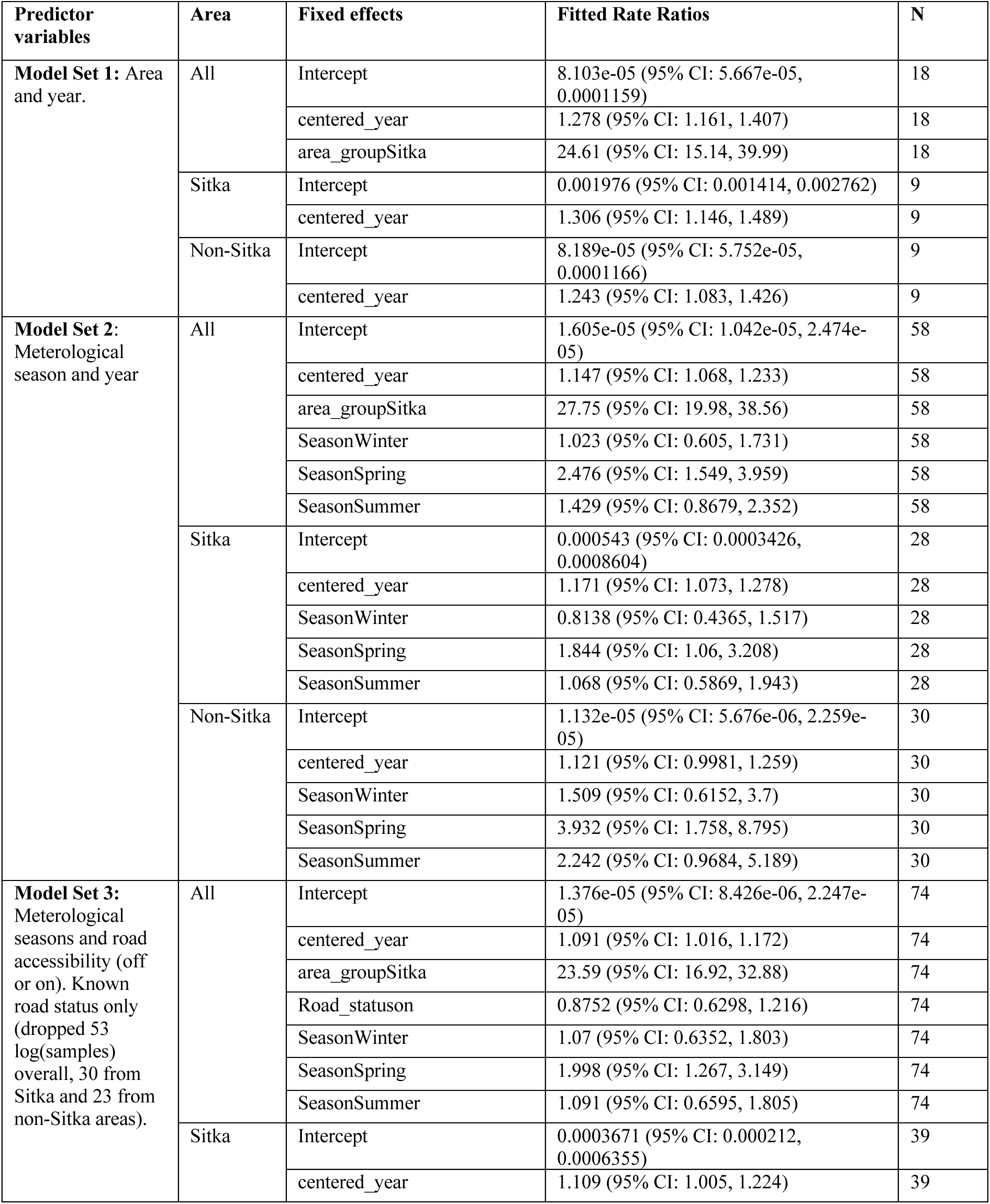
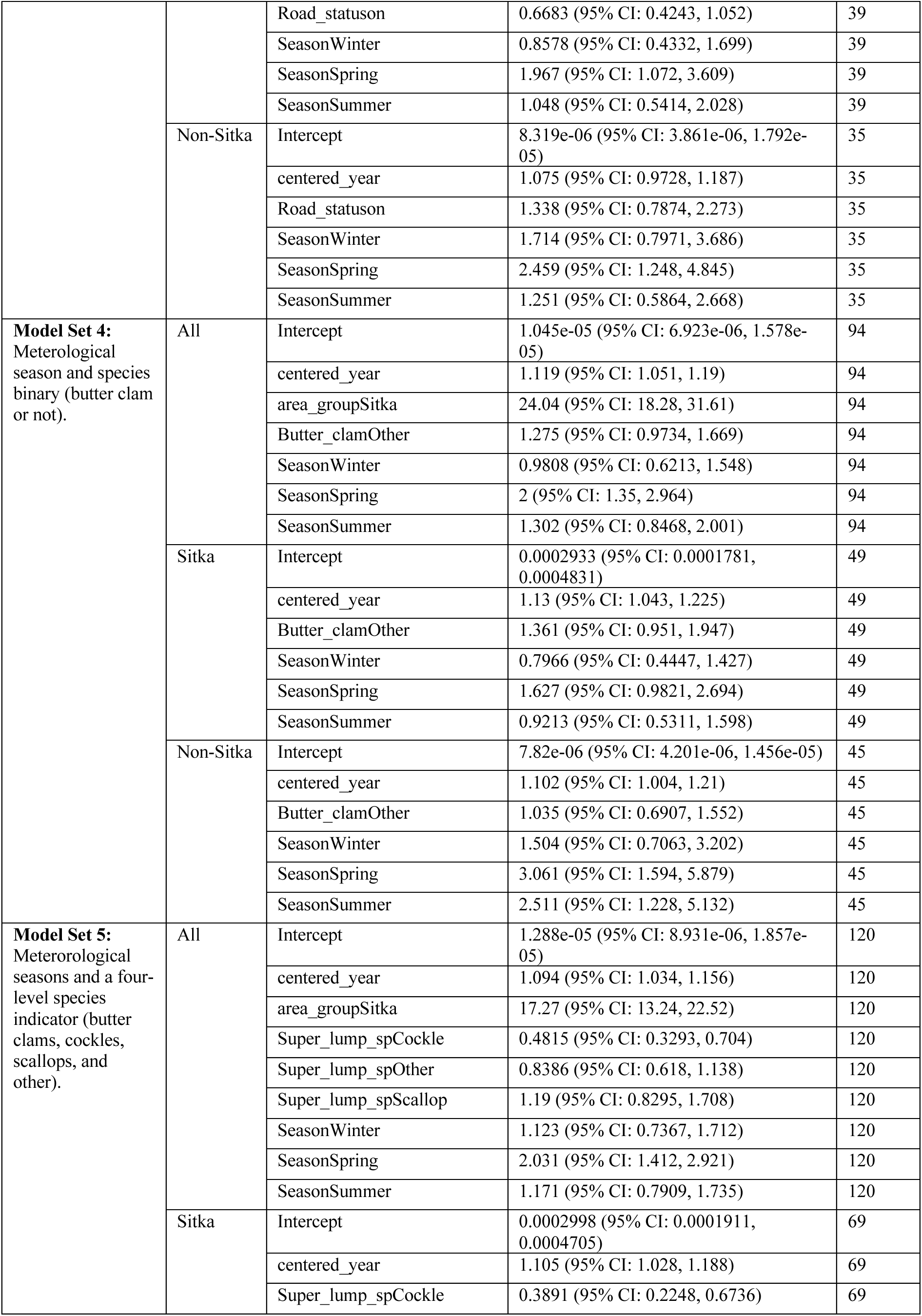
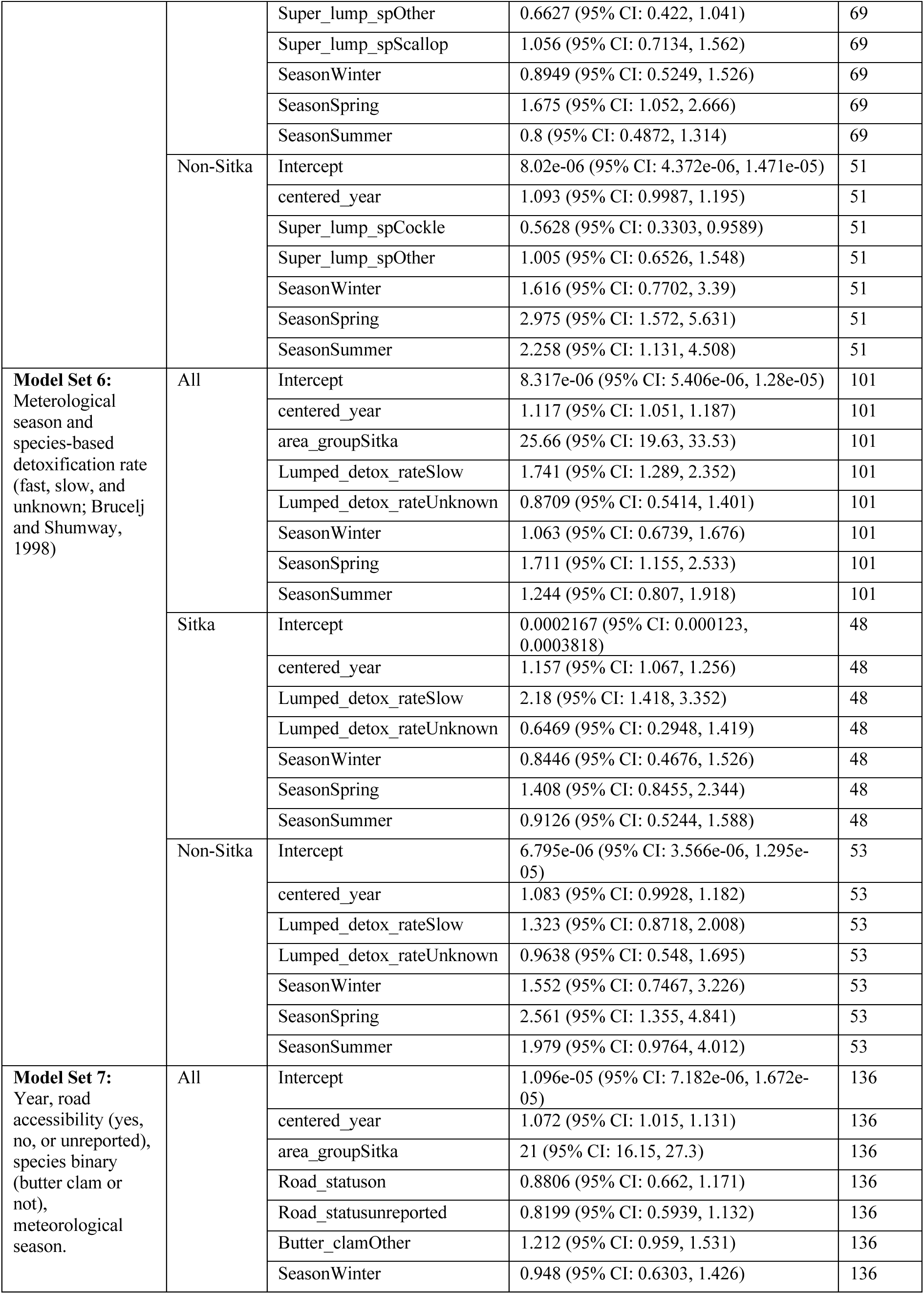
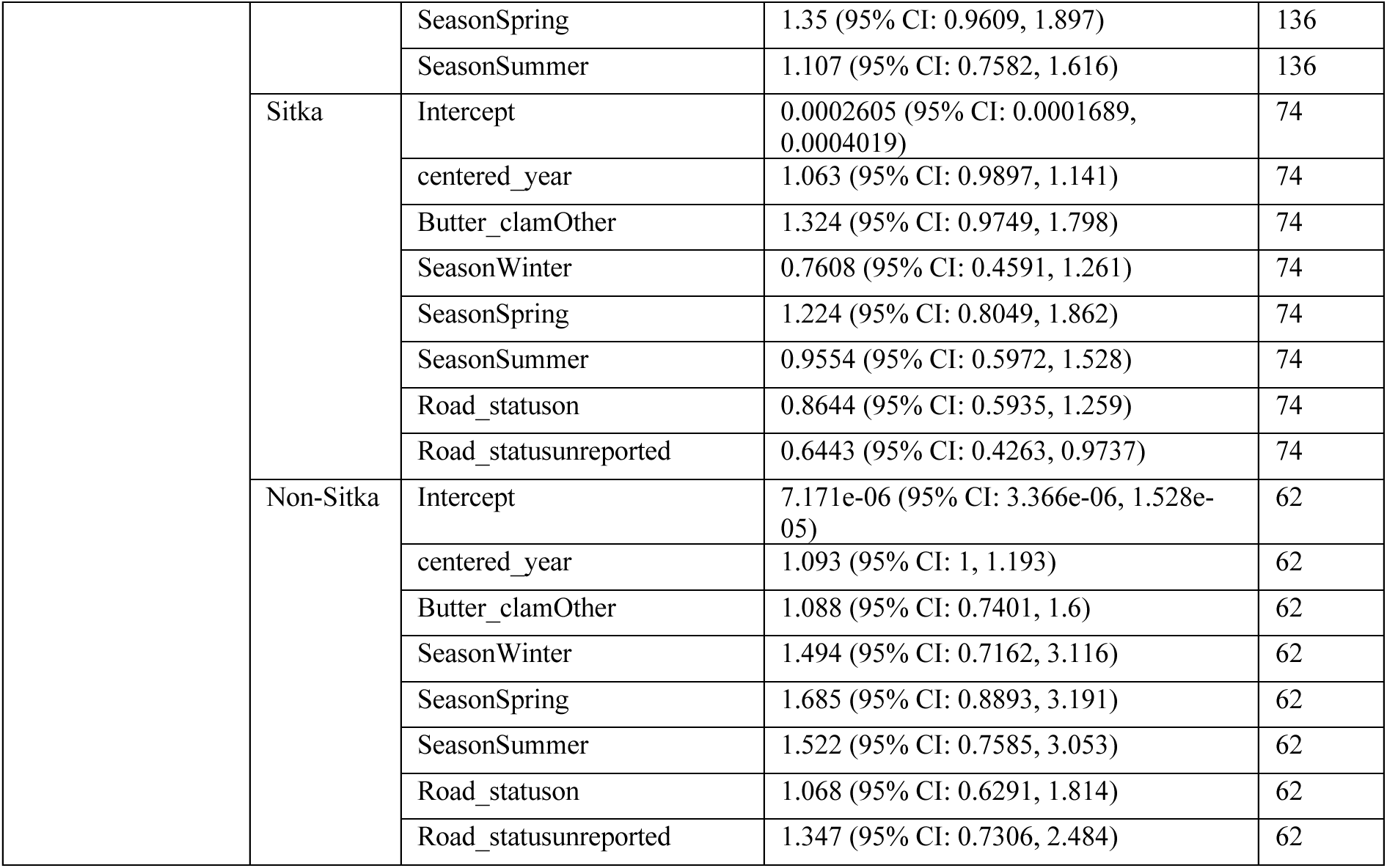
Fitted model rate ratios (on the response scale) for all fixed effects for per capita toxin testing rates. Each "model set" includes a model with Sitka vs. non-Sitka as a fixed effect, then two separate models analyzing only Sitka or non-Sitka data.

We coded all data manipulation and regressions in R, with regressions fit using the glm.nb function of the lme4 package (43). We assessed all model fits by examining the Pearson residuals in QQ plots, histograms, and against predictors using the R package ggResidpanel (44).

## 3. RESULTS

### 3.1 Temporal trends

Overall, the number of samples that community members sent for toxin testing has increased since the start of the program, though with a large decrease from 2021 to 2022 following shortages in toxin testing reference materials during the COVID-19 pandemic that prevented STEARL from conducting testing (Figure 2; Table 2). However, this material shortage and limited testing was followed by the largest number of samples sent by community members in 2024. In Model Set 1, with only year as a predictor, the per capita rate of testing increased 28% per year on average (TR = 1.28, 95% CI: 1.161, 1.407; Table 3). The rate ratio associated with year shrank with the inclusion of additional predictor variables, but was always positive. Whether examined in isolation, as in Model Set 1, or conditioning on season, species, or road accessibility, the effect of year did not vary significantly between Sitka and non-Sitka areas, with estimated effects from isolated Sitka and non-Sitka models within error across all model sets (Table 3).

**Figure 2:**
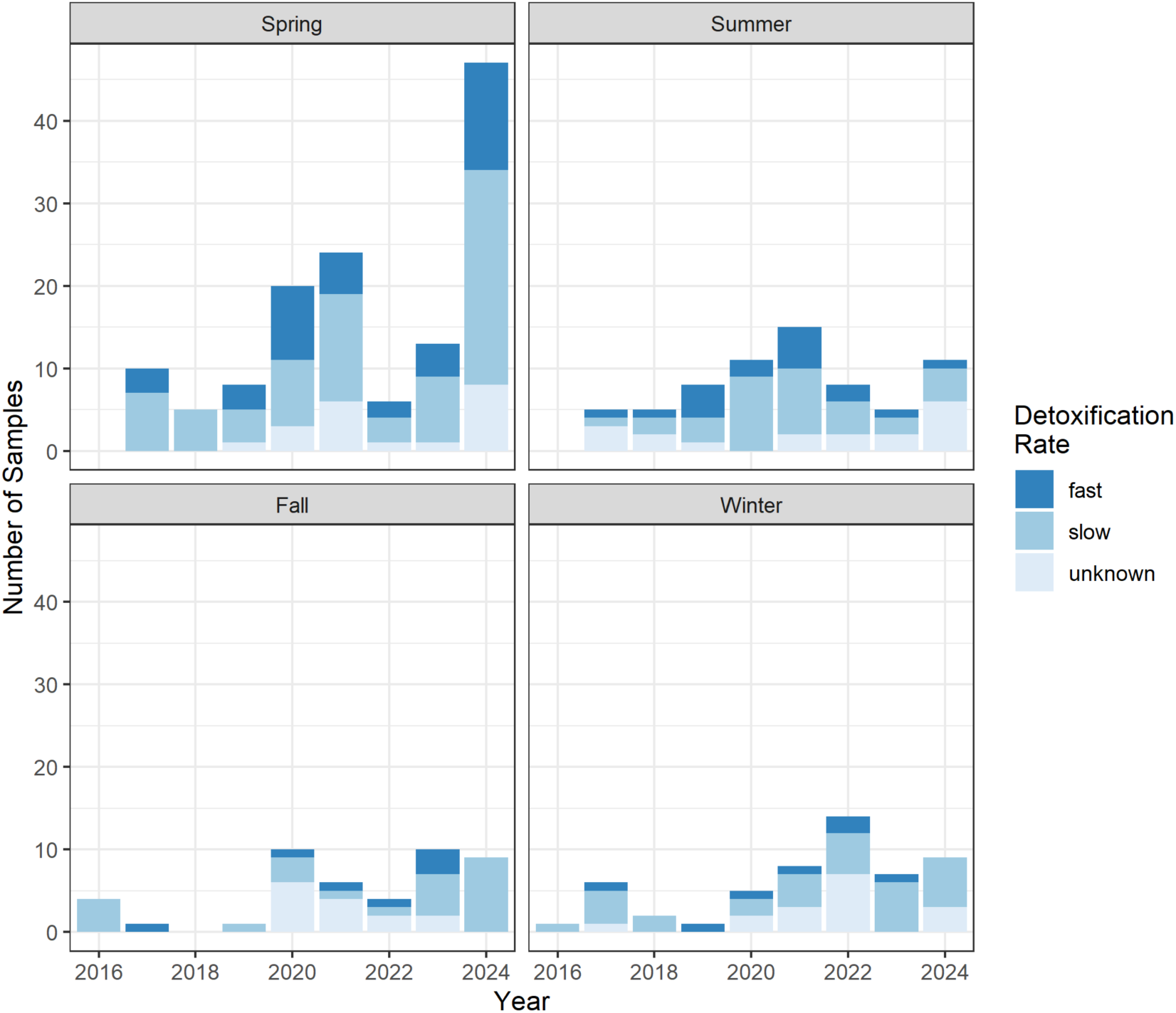
The number of shellfish samples sent to the Sitka Tribe of Alaska Environmental Research Lab (STAERL) by individual harvesters over time, split by season and species-based detoxification rate. Spring runs from March through May, summer from June through August, fall from September through November, and winter from December through February. Detoxification rates are according to Bricelj and Shumway (1998).

### 3.2 Seasonal trends

Shellfish testing utilization varied by season. Spring had the highest number of tested samples in most years and overall, followed by summer, winter, and then fall (Table 2; Figure 2). When only considering season and year, as with Model Set 2, the fitted rate ratio from fall to spring was 2.48 (95% CI: 1.549, 3.959), decreasing to near unity for the fall to winter rate ratio (Table 3). The increase in sampling rates from fall to spring are stronger in non-Sitka areas than in Sitka, with non-Sitka areas also displaying a lesser, but consistent estimated increase from fall to summer sampling rates. When conditioning on road status or species-related variables, spring and summer seasons are still typically associated with large increases in sampling rates relative to fall in most models in non-Sitka areas. In Sitka, however, only spring is consistently associated with an increase in testing rates relative to fall and even then, the effect is rarely significant.

### 3.3 Spatial and accessibility trends

Our hypothesis that larger physical barriers to harvest sites or testing, such as lack of roadside access to harvesting sites or needing to ship samples by air to STAERL, would reduce testing utilization was only slightly supported. Across all fitted models, the spatial effect of Sitka vs. non-Sitka emerged as having the largest effect on the number of shellfish samples sent in for testing (Table 3), with the estimated effect size ranging from 17 to 27 depending on the additional included parameters. This effect size was an order of magnitude larger than any other estimated effect size, though we note that the lumping of all non-Sitka areas could elide substantial heterogeneity across non-Sitka communities. The “Sitka effect” is also apparent in the Sitka and non-Sitka submodels, where the former have an intercept term an order of magnitude larger than the latter, again highlighting that the modeled per-person testing rate is approximately 20 times higher in Sitka than outside of it.

In contrast to harvesting location, road access was not a significant predictor of shellfish testing rates and had opposed, though highly uncertain, estimated effects in Sitka and non-Sitka areas in Model Set 3 (Table 3). In Sitka, being on the road system was associated with reduced sampling rates, while outside of Sitka the estimated effect was weakly positive, though in both cases the uncertainty suggested no meaningful effect. We found no evidence to support the inclusion of interaction terms between road status and season, suggesting that seasonally favorable weather does not significantly alter the importance of road accessibility. As previously noted in Section 3.1, the samples with “unreported” road status dropped from Model Set 3 were highly unbalanced seasonally. It is possible that they are also not missing at random with respect to road status.

### 3.4 Species and detoxification rate trends

Our hypothesis that testing utilization rates would be highest in spring and summer–the warmer months and riskier period according to the adage of only harvesting in months that contain the letter “r”–also has implications for variations we hypothesize in terms of species detoxification rates. We anticipate that greater risks in warmer spring months will lead harvesters to test faster toxifiers/detoxifiers such as mussels, cockles, and littleneck clams more in the spring. However, contrary to this hypothesis, we find no support for seasonally specific changes to species harvest rates as the inclusion of interaction terms between species variables or detoxification rates and seasons was not supported by likelihood ratio tests across all model sets and location-specific submodels. Nevertheless, we do find that species based detoxification rates were strongly associated with increased shellfish testing rates for slow detoxifiers - a category that includes both butter clams and scallops, the two most commonly submitted species - relative to fast detoxifiers, whereas fast detoxifying species did not have differentiated testing rates from those with unknown detoxification rates. Butter clams are the most commonly tested species across all years and seasons, comprising 44% of all submitted samples, 42% of those from Sitka, and 46% of those from outside of Sitka. Since butter clams comprise nearly half of the submitted samples, the “not a butter clam” variable tested in Model Set 3 had an estimated rate ratio near one overall and outside of Sitka, and a moderately positive rate ratio in Sitka (Table 3). When splitting the data into the three most common species and an “other” category, as in Model Set 4, only the rate ratios for cockles were strongly predictive relative to butter clams, with estimated rate ratios near 0.5 such that cockles are half as likely to be submitted for testing as butter clams. Though scallops comprise only 18% of submitted samples across all data, all scallops come from Sitka, where they are 42% of the submitted samples and comparable to butter clams in numbers, so the scallop species predictor was confounded with the area predictor in these species models.

## 4. DISCUSSION

This study evaluated the numbers of shellfish samples that members of the public sent to a tribally run shellfish toxin testing lab to be tested for toxins from 2016-2024 and across participating communities in the Gulf of Alaska. The goal was to identify temporal, spatial, and species patterns to evaluate possible explanations for observed heterogeneity and better understand usage. We were particularly interested in elucidating possible influences of risk perceptions and of harvesting and testing accessibility. Research has suggested that difficulty accessing shellfish reduces community harvesting and consumption (11). With respect to harvester risk perception, we anticipated that shellfish harvesting and testing rates might vary seasonally by species given that the slow detoxification rates of butter clams, in particular, are well publicized by the SEATOR consortium and other Gulf of Alaska research groups and that species differences in detoxification rates is known to harvesters (11,15,45). Our analysis finds that, overall, the number of samples that community members sent for toxin testing has increased through time, despite a drop in samples tested during the COVID-19 pandemic. The increase in utilization through time may relate to factors including greater awareness, familiarity, buy-in, and trust in the program, may be partially explained by the switch from charging for sample testing to free access in 2021, or may relate to greater overall rates of shellfish harvesting since the SEATOR regular monitoring program has grown. Since the inception of this program and the public testing option there has not been a single reported incident of PSP in Southeast Alaska, compared to more than 50 cases of PSP in Southeast Alaska from 2000-2016 (12). Contrary to our expectations, we found mixed support for increased testing in response to more convenient shellfish or testing access or in response to species detoxification rates, and we found no support for seasonal modulation of the effects of either species or convenience.

Seasonal differences in testing utilization were fairly consistent across locations and years; in both Sitka and non-Sitka areas, spring had the highest utilization, and fall and winter had the lowest testing utilization. Spring is an important traditional harvest season for clams (3,46,47). While winter months are traditionally known as safer for shellfish harvest (15,17), September, March, and April are the only non-summer months with sufficiently low tides during daylight hours, increasing shellfish accessibility. It is unclear from our data whether lower winter and fall testing rates reflect lower rates of shellfish harvest, which would be unexpected but could reflect the nighttime low tides and poor weather common to both colder seasons, or lower perceptions of risk during fall and winter months such that harvesters do not feel the need to test shellfish prior to consumption. Given the high documented harvest of shellfish, especially on Kodiak Island and in smaller Southeast Alaskan communities (32), it is unlikely that testing utilization patterns mirror shellfish harvesting patterns. Rather, the convenience factor of being in Sitka may encourage local harvesters to test year-round, reducing the differences in testing rates between seasons, while outside of Sitka testing rates may be more responsive to seasonal perceptions of risk.

We were surprised by our finding that road accessibility of harvesting sites does not influence shellfish testing rates. Relative to the barrier of shipping samples to Sitka, road accessibility of shellfish may be a trivial concern. Furthermore, harvesters may preferentially seek shellfish off the road system to reduce risks of urban shellfish contaminants like fecal coliforms or oil. Surveys of recreational shellfish harvesters in Washington and France found that they would be willing to pay hundreds of dollars annually to avoid pollution concerns at their preferred harvesting sites (48,49); in rural Alaska, the value of a pristine harvesting site may be more associated with beaches that are boat access only, avoiding point-based pollution sources in and around community centers such as harbors, wastewater treatment sites, and road runoff. The lack of strong evidence for any interaction between season and road accessibility is also slightly surprising, as colder months bring potentially dangerous ocean conditions. The strong seasonal biases of shellfish samples without known road status (skewed toward fall in Sitka and strongly toward summer and spring outside of Sitka) might have masked or altered the importance of seasonal interactions with road status.

In contrast to road accessibility, the strength of the “Sitka” effect, where per capita testing rates were much higher in Sitka than outside of it, does suggest that our hypothesis that convenience increases sampling rates was partially correct. As a per capita measure, the strength of this effect is sensitive to the populations of interest. In the case of our “non-Sitka” group, this population is quite high as it includes the full Kenai Peninsula Borough and full Nome Census Area populations despite only two shellfish samples originating from either location. The included boroughs differ substantially in their demographics and reported reliance on shellfish harvests, with some such as Haines reporting low shellfish use, while others such as Hoonah report higher use (32,42). Using the “harvesting population” of a given borough would be a better metric for testing service utilization rates, but this is not easily determined on a borough basis from the available harvesting data (32). The effect of Sitka on shellfish harvesting rates is strongly positive even with large changes to the “non-Sitka” population, but we do acknowledge that within the non-Sitka communities, there are likely large differences in testing utilization rates.

We were surprised to not see direct evidence for seasonal and species relationships consistent with detoxification rate-based risk avoidance in harvesters. The differential detoxification rates between butter clams and other commonly harvested shellfish species are known (15,50) and have been repeatedly emphasized in SEATOR communications and, thus, we had anticipated a season-species interaction. Instead, we could not reject the null hypothesis that there are no seasonally modulated changes to species testing rates. While there were low sample sizes to test this interaction (for Model Set 6, n = 101, *X*2 = 3.16, p =0.53), it is also possible that harvester preferences for slowly detoxifying species outweighs the perceived seasonal changes in risk. Butter clams are the most commonly tested species across all seasons, likely reflecting their desirableness in subsistence and recreational harvest (22,32), followed by similarly slowly detoxifying scallops. Since butter clams and scallops were so commonly tested, we did find that our “slow detoxification” predictor was associated with substantial increases in the expected number of tested samples, particularly in Sitka. However, if detoxification rates rather than species preferences were driving that trend, we would expect the “fast detoxifying” group of cockles, littleneck clams, mussels, and oysters to be differentiated from the group of unknown detoxifiers, or for species testing preferences to vary significantly across seasons. It is possible that if we were able to normalize shellfish testing rates by true harvesting rates, testing rates might be elevated relative to harvesting rates in those species considered higher risk.

Our findings on service utilization for individual shellfish testing suggest several future lines of inquiry, as well as a few caveats for our conclusions. Since we did not have access to harvester information, we cannot comment or speculate on factors that make individuals more likely to send in samples, including risk perception and accessibility explicitly. Additionally, one sampler sending in repeated shellfish tests is indistinguishable in our dataset from multiple harvesters sending in one sample each, but the scenarios suggest significantly different factors influencing service utilization. We also cannot comment on the relationship between a community’s regular testing program and individual testing; community advisories from the regular SEATOR monitoring program could encourage harvesters to test their personal samples, eliminate the perceived need for additional testing, or discourage harvesting altogether. While we grouped all non-Sitka boroughs into a single category for analysis, the number of samples sent per borough suggests high variance in the testing utilization rates amongst communities with high reliance on shellfish harvests (32); communities with lower testing rates may have less perceived or historical risk, or may have regular monitoring of the most popular beaches. Finally, SEATOR partners in some communities may directly facilitate individual shellfish harvesting by taking care of shipping logistics, while in communities outside the SEATOR consortium this logistical piggybacking is not possible. We encourage future interview-based and population-based studies to directly elucidate the relationship between risk perceptions, testing and site accessibility, harvesting rates, and toxin testing rates.

This study has several important implications for reducing shellfish harvesting risks in Alaska and in other subsistence harvesting communities. The consistency in tested shellfish species, regardless of season, suggests that risk awareness communications focusing on choosing different species may have less utility for harvesters, who are searching for ways to reduce their risks given their a priori choice of preferred species. The fact that spring is the most popular season for shellfish testing is encouraging as the harmful algal bloom season expands into months previously considered low risk like March and April, though we note that increased testing rates may not closely track overall shellfish harvesting rates. Since regular SEATOR testing began in 2016, April has already hosted 19 shellfish samples from five different communities above FDA toxin limits from active harmful algal blooms, including one from Sitka on April 8th of 2025 (51). As expanding bloom windows increase the probability for HAB events even in seasons and locations where they previously did not occur, regular monitoring programs and individual testing programs such as this strengthen the capacity for harvesters to make safe choices about their harvest and food security.

## 5. CONCLUSION

Utilization rates of a tribally led shellfish toxin testing program have grown across the Gulf of Alaska since 2016, a period over which there have been no reported cases of shellfish toxin related diseases in Southeast Alaska. While tribal governments in the region regularly monitor specific shellfish harvesting beaches for toxins, individual community members harvesting from unmonitored locations are also encouraged to test their shellfish before consumption. The consistency of species and seasonal testing choices by individual harvesters, in spite of cyclical changes in toxin risk and off-road site accessibility suggests that testing practices are strongly influenced by personal preference for specific sites or species. By developing a better understanding of the influences of risk perception and access to adaptation resources on participation rates in risk reduction strategies like shellfish toxin testing, environmental risk managers can better predict and respond to changes in demographics, environmental conditions, and harvest and utilization patterns and better tailor risk communication and community support.

## 6. DECLARATIONS

### 6.1 Ethics approval and consent to participate

Not applicable.

### 6.2 Consent for publication

Not applicable.

### 6.3 Availability of data and materials

The dataset supporting the conclusions of this article is available in the Dryad repository at https://doi.org/10.5061/dryad.dfn2z35dr.

### 6.6 Authors’ contributions

EGK led the formal analysis and manuscript writing. JRH contributed to conceptualization, methodology, manuscript review, and funding acquisition. SC and KL contributed to investigation and data curation. JF and CW contributed to resources and funding acquisition. MOG and HBR contributed to conceptualization, methodology, manuscript review, and funding acquisition. All authors reviewed the final manuscript.

### 6.7 Acknowledgements

Not applicable.

